# Effects of social support on depression risk during the COVID-19 pandemic: What support types and for whom?

**DOI:** 10.1101/2022.05.15.22274976

**Authors:** Karmel W. Choi, Younga H. Lee, Zhaowen Liu, Daniel Fatori, Joshua R. Bauermeister, Rebecca A. Luh, Cheryl R. Clark, André R. Brunoni, Sarah Bauermeister, Jordan W. Smoller

## Abstract

**Background:** Rates of depression have increased worldwide during the COVID-19 pandemic. One known protective factor for depression is social support, but more work is needed to quantify the extent to which social support could reduce depression risk during a global crisis, and specifically to identify which types of support are most helpful, and who might benefit most.

**Methods:** Data were obtained from participants in the *All of Us* Research Program who responded to the <u>CO</u>VID-19 <u>P</u>articipant <u>E</u>xperience (COPE) survey administered monthly from May 2020 to July 2020 (N=69,066, 66% female). Social support was assessed using 10 items measuring emotional/informational support (e.g., someone to confide in or talk to about yourself or your problems), positive social interaction support (e.g., someone to do things with to help you get your mind off things), and tangible support (e.g., someone to help with daily chores if sick). Elevated depression symptoms were defined based on having a moderate-to-severe (≥10) score on the Patient Health Questionnaire (PHQ-9). Mixed-effects logistic regression models were used to test associations across time between overall social support and its subtypes with depression, adjusting for age, sex, race, ethnicity, and socioeconomic factors. We then assessed interactions between social support and potential effect modifiers: age, sex, pre-pandemic mood disorder, and pandemic-related stressors (e.g., financial insecurity).

**Results:** Approximately 16% of the sample experienced elevated depressive symptoms. Overall social support was associated with significantly reduced odds of depression (adjusted odds ratio, aOR [95% CI]=0.44 [0.42-0.45]). Among subtypes, emotional/informational support (aOR=0.42 [0.41-0.43]) and positive social interactions (aOR=0.43 [0.41-0.44]) showed the largest protective associations with depression, followed by tangible support (aOR=0.63 [0.61-0.65]). Sex, age, and pandemic-related financial stressors were statistically significant modifiers of the association between social support and depression.

**Conclusions:** Individuals reporting higher levels of social support were at reduced risk of depression during the early COVID-19 pandemic. The perceived availability of emotional support and positive social interactions, more so than tangible support, was key. Individuals more vulnerable to depression (e.g., women, younger individuals, and those experiencing financial stressors) may particularly benefit from enhanced social support, supporting a precision prevention approach.

## Introduction

Rates of depression have increased worldwide during the COVID-19 pandemic (1), highlighting the importance of identifying modifiable factors for targeted approaches to prevention and intervention during a global crisis. Social support is a robust protective factor for depression, both in general (2) and during times of stress (3), demonstrating promise for reducing the population-level burden of depression during a highly stressful pandemic. Accordingly, a growing number of studies on the overall relationship between social support and mental health during the COVID-19 pandemic, have linked greater social support to reduced risk for depression and other negative mental health outcomes (4–6).

However, previous studies have been limited for several reasons. First, many have relied on relatively modest samples that were not followed longitudinally. Second, studies of elevated depressive symptoms during the COVID-19 pandemic have not consistently explored subtypes of social support, which include tangible supports characterized by the availability of instrumental help for everyday or crisis needs; emotional supports characterized by the availability of a listening or confiding ear; informational supports characterized by the availability of advice or knowledge from others; and positive social interactions (7). For instance, during a pandemic, tangible support may become more emotionally salient than other forms of support, due to the risk of illness, contagion, and other daily disruptions, while the relevance of positive social interactions may be diminished in the context of pervasive social distancing. Understanding which type(s) of social support most influence depression risk during the pandemic could highlight which aspects should be addressed in both individual- and population-level interventions. Third, it is still unclear which groups may benefit from increased social support. For example, the protective role of social support may vary by intrinsic factors such as age and sex, which may influence both depression risk and how social support impacts this risk, or by pre-pandemic factors such as prior mental health histories, which may increase vulnerability to subsequent distress. Additionally, risk factors may emerge during the pandemic such as financial stressors (e.g., loss of income), which either limit one’s ability to benefit from social support or indicate where social supports are particularly needed. A better understanding of potential effect modifiers, across a range of intrinsic, pre-pandemic, and during-pandemic characteristics, could provide more insight into where interventions aimed at enhancing social support could have the largest impact. Knowledge, for example, about whether pre-existing or concurrent risk factors are more salient during the pandemic could inform the targeting of such interventions.

More detailed work in large prospective cohorts is needed to quantify the extent to which social support may promote resilience to elevated depression symptoms during the COVID-19 pandemic, and specifically, to identify which types of support may prove most helpful, and who might benefit most. One opportunity to rigorously investigate these questions is within the *All of Us* Research Program, an ongoing, diverse US nationwide research cohort where a longitudinal survey focused on mental health, coping, and other experiences including social support was administered in the first year of the pandemic. Using longitudinal data across the first three survey waves completed by *All of Us* Research Program participants (N=69,066), we tested associations between perceived social support and its subtypes with depression risk, and assessed potential effect modification by intrinsic (e.g., sex, age), pre-pandemic (e.g., prior mood disorder), and pandemic-related (e.g., financial stress) risk factors.

## Methods

### Cohort description

The *All of Us* Research Program (AoU) (8) has enrolled more than 482,000 participants as of April 2022. More than 80% of participants are from communities that have been underrepresented in biomedical research based on the following characteristics: race and ethnicity; age; sexual orientation and gender identity; low income and educational attainment, rural residence; and disability. The Institutional Review Board of the *All of Us* Research Program has approved all study procedures, and participants provided informed consent to share electronic health records (EHRs), surveys, and other study data with qualified investigators for broad-based research.

### Study sample

The <u>CO</u>VID-19 <u>P</u>articipant <u>E</u>xperience (COPE) survey was administered electronically to AoU participants to assess the longitudinal impact of the pandemic and included questions on COVID-19 symptoms, physical and mental health, social distancing, economic impacts, and coping strategies. The first three waves of the COPE survey (administered in May, June, and July of 2020) included assessments of social support and depressive symptoms and are the focus of the current study. A total of 69,066 respondents completed the COPE survey at least once across these three timepoints.

It has been previously reported that research volunteers tend to be healthier and have higher socioeconomic status compared to the underlying source population (9). In recent prior work, we found some evidence of “healthy volunteer bias” among the COPE survey participants and demonstrated the utility of inverse probability weighting in offsetting potential bias (10). We thus calculated inverse probability weights (IPWs) for COPE survey completion at each timepoint using sex assigned at birth, self-reported race and ethnicity, birthplace, educational attainment, marital/partnership status, health insurance status, employment status, homeownership, and current age as predictors, to use as weights in the primary analyses.

### Social support

Social support (received in the past month) was measured using 10 items from the RAND Medical Outcome Study (MOS) Social Support Survey Instrument (7), using a five-point scale ranging from *1 = None of the time* to *5 = All the time*. These items can be classified into subtypes of tangible support, emotional/informational support, and positive social interaction support (7) (for a list of all items and their corresponding subtypes, see **Supplementary Table 1**). After excluding participants who were missing responses for all 10 social support items at each wave (N=570 in May, N=270 in June, N=480 in July), an overall social support score was calculated as the mean rating across all completed items, and scores for each support subtype were calculated as the mean rating across all completed items for a given subtype. All scores were standardized (mean=0, SD=1) prior to analysis to facilitate interpretation. Correlations between each subtype ranged from r=0.65 to 0.83 (**Supplementary Table 2**), especially between emotional/informational support and positive social interaction.

To further characterize the potential impact of social support subtypes and their combinations, we derived dichotomous indicators for individuals who reported above-versus below-average (including average) levels of each support subtype. We then created an eight-level categorical exposure variable to indicate individuals who endorsed higher levels on either all three support subtypes, only two subtypes, only one subtype, or no subtypes.

### Depression

Depressive symptoms were assessed using the Patient Health Questionnaire (PHQ)-9 (11), a nine-item checklist that assesses symptoms of depression in the past two weeks using a four-point scale response format (ranging from *0 = Not at all* to *3 = Nearly every day*). The scale is scored by summing responses to all nine items, with total scores ranging from 0 to 27. Total scores of 5, 10, and 20 represent validated cut-points for mild, moderate, and severe depression (11,12); scores in the moderate-to-severe range of 10 or above thus reflect clinically elevated symptoms of depression (hereafter, referred to as “depression” to be concise).

### Covariates

We used linked EHR data to establish whether participants had a pre-pandemic history of mood disorder diagnosis at any time prior to January 21^st^, 2020, the date of the first reported COVID-19 case in the United States (13), defined by two or more qualifying diagnostic codes mapped to “Mood disorder” code 46206005 in the Systematized Nomenclature of Medicine (SNOMED; a common coding scheme for harmonizing different coding vocabularies or ICD versions across health systems). Those without qualifying codes or, to be conservative, did not have linked EHR data were considered not to meet criteria for such a history. Linked EHR data were available for 54,426 participants (79%). Participants also responded to the COPE survey about past-month potential financial stressors, including not having enough money to pay for housing, gas/fuel, food, medications, or not having a regular place to sleep or stay. Given that COPE survey respondents generally did not endorse many financial stressors, we binarized this variable to indicate the presence of at least one stressor. We also extracted information from the AoU baseline survey on sex assigned at birth, current age, homeownership, employment status, educational attainment, health insurance status, and self-reported race and ethnicity.

### Statistical analysis

Although the COPE survey was administered multiple times over the course of the pandemic, not all participants completed each assessment. To accommodate both missingness and within-subject correlations across survey measurements (14), we first fitted IPW-adjusted, mixed-effects logistic regression models using the *lme4* R package to determine the time-varying relationships between social support and depression with subject-specific random intercepts and fixed effects for the three survey timepoints (i.e., May, June, and July of 2020). We adjusted for potential confounding factors including sex, age, homeownership, employment status, educational attainment, health insurance status, and self-reported race and ethnicity in models testing main effects of overall social support and specific subtypes of social support. Second, we similarly fitted a mixed-effects model to test the association between the categorical variable of support subtype combinations and depression, using the category with lower support on all three subtypes as the reference group. Third, we assessed binary factors—sex assigned at birth (female versus male), current age (below versus above 65), pre-pandemic mood disorder diagnosis (any versus none), and COVID-related financial stress (any versus none)— as potential modifiers of the association between social support and depression during the pandemic. As a sensitivity analysis, we also fitted logistic regression models testing lagged associations between overall social support at baseline and subsequent depression at the second wave only (one-month window) or at the second or third survey wave (two-month window), excluding individuals with elevated depressive symptoms at baseline.

All analyses were performed using data from the *All of Us Registered Tier Version R2020Q4R2* on the AoU Researcher Workbench (https://workbench.researchallofus.org), a cloud-based platform where approved researchers can access and analyze data (15). We used R version 4.1.0 in a Jupyter Notebook contained in the AoU Workbench to query data, perform statistical analyses, and generate tables and figures.

## Results

Sample characteristics are summarized in **Table 1**. Participants were predominantly female (66%) and self-reported White (82%), with an average age of 59 years (SD=16), with 8% reporting any pandemic-related financial stressors. On average, 16% of study participants met criteria for clinically elevated (i.e., moderate-to-severe) symptoms of depression across the survey months of May, June, and July, and 14% of the sample were identified to have a pre-pandemic mood disorder diagnosis based on linked EHR data.

**Table 1.** Sociodemographic characteristics of the 69,066 COPE survey respondents in the All of Us Research Program.

| **Categorical variables** | **N** | **Percent** |
| --- | --- | --- |
| Sex assigned at birth |  |  |
| Male | 23,598 | 34% |
| Female | 45,468 | 66% |
| Self-reported race |  |  |
| Asian | 2,036 | 3% |
| Black or African American | 4,082 | 6% |
| White | 56,424 | 82% |
| I prefer not to answer/None indicated/Skipped | 4,410 | 6% |
| Another single population/More than one population/None of these | 2,114 | 3% |
| Self-reported Hispanic ethnicity | 4,879 | 7% |
| Birthplace |  |  |
| Outside of the U.S. | 6,766 | 10% |
| U.S. | 62,300 | 90% |
| Educational attainment |  |  |
| Less than college | 5,866 | 8% |
| College or beyond | 63,200 | 92% |
| Employed | 37,784 | 55% |
| Has health insurance | 67,172 | 97% |
| Has homeownership | 47,710 | 69% |
| Married/partnered | 43,723 | 63% |
| Experienced any COVID-19 symptom(s) | 2,276 | 3% |
| Had any mood disorder diagnosis prior to COVID (lifetime) | 9,930 | 14% |
| Had any financial stress due to COVID | 5,181 | 8% |
| **Continuous variables** |  |  |
| Current age | 58.9 | 16.0 |
| Income | 90893.3 | 59357.6 |

After adjusting for sociodemographic and clinical factors, overall social support was inversely associated with depression (aOR [95%CI]=0.44 [0.42-0.45], p<2.0×10^−16^) (**Table 2**). As shown in **Figure 1a**, the predicted probabilities of depression were elevated at low levels of reported social support and declined towards zero at the highest levels of social support. This was consistent with lagged sensitivity analyses, where overall social support at baseline was prospectively associated with depression in the subsequent one- or two-month windows (aORs 0.67-0.68; **Supplementary Table 5**), even after excluding individuals with elevated symptoms at baseline.

**Table 2.**
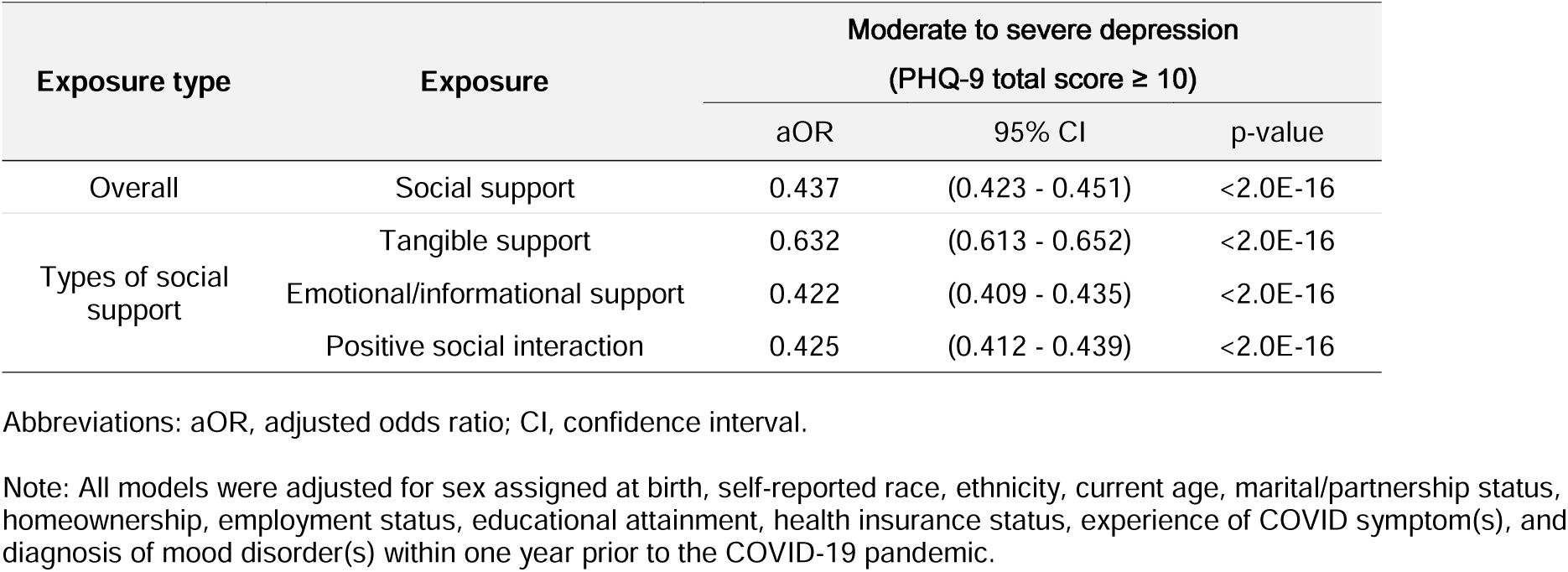
Inverse-probability weighted mixed-effects logistic regression analysis examining the effects of social support on depression, and variations by types of social support.

**Figure 1.**
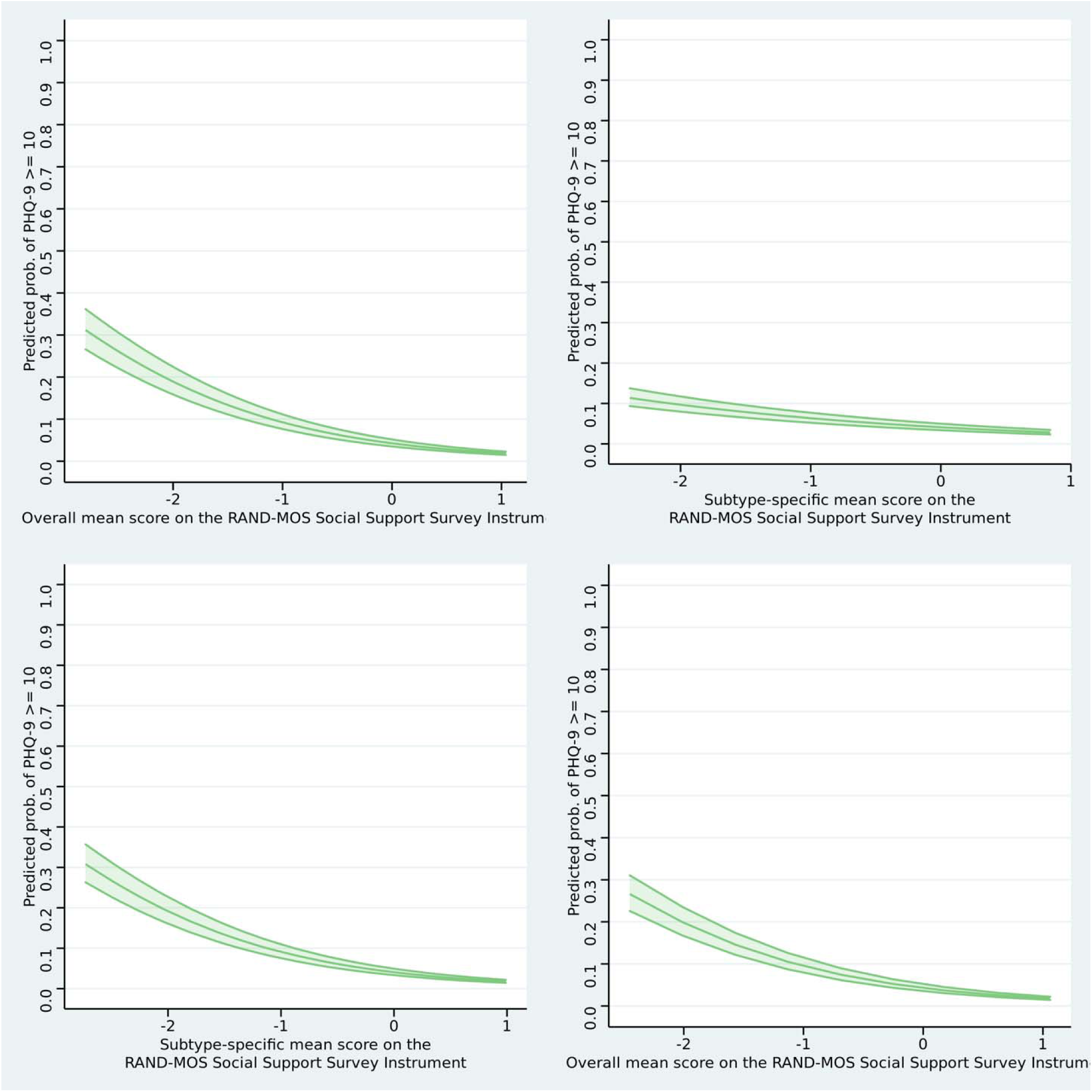
Predicted probabilities of moderate-severe depressive symptoms at different levels of overall and subtype-specific social support (standardized) : (a) overall social support (top left), tangible support (top right), (c) emotional/informational support (bottom left), (d) positive social interaction (bottom right).

The emotional/informational support subtype (**Figure 1c**) showed the largest inverse association with depression (aOR [95%CI]=0.42 [0.41-0.43], p<2.0×10^−16^), followed by positive social interaction (**Figure 1d**) (aOR [95%CI]=0.43 [0.41-0.44], p<2.0×10^−16^) (**Table 2**). Tangible support (**Figure 1b**) was also associated with a smaller but nonetheless significant reduction in depression odds (aOR [95%CI]=0.63 [0.61-0.65], p<2.0×10^−16^). When examining combinations of social support subtypes, a marked dose-response gradient (**Figure 3; Table 3**) was observed. Compared to those reporting lower support on all subtypes, those with higher tangible support alone showed a modest reduction in the odds of depression (aOR [95% CI]=0.89 [0.81-0.98]) compared to those with higher positive social interaction (aOR [95% CI]=0.43 [0.35-0.51]) or emotional/informational support (aOR [95% CI]=0.38 [0.34-0.44]) alone. Endorsing higher levels of at least two subtypes of support was linked to greater reductions in the odds of depression, with a particularly strong protective association among those endorsing both emotional/informational support and positive social interaction (aOR [95% CI]=0.22 [0.19-0.25]). However, endorsing higher levels of all three subtypes of support appeared most protective (aOR [95% CI]=0.15 [0.14-0.16]).

**Table 3.**
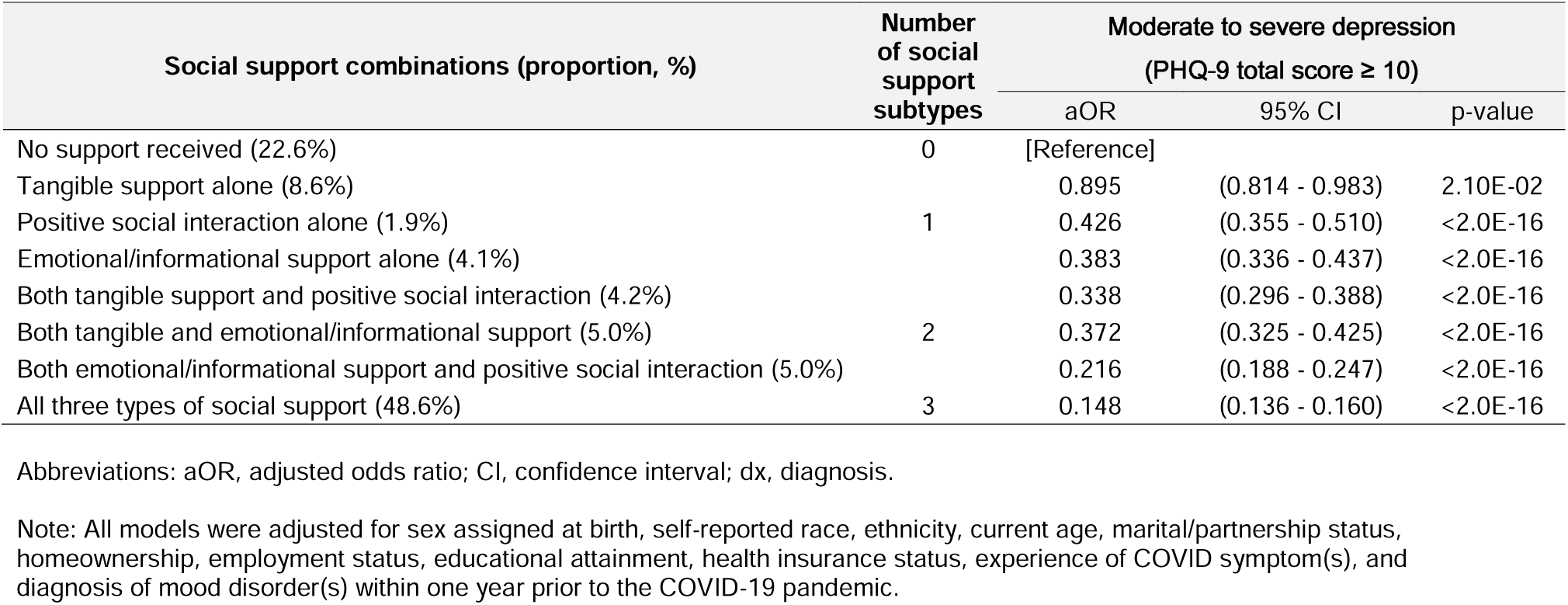
Inverse-probability weighted mixed-effects logistic regression analysis examining the effects of specific combinations of social support types on depression.

Lastly, we identified overall effect modification by sex (β=-0.075, interaction *p*=0.017), age (β=0.10, interaction *p*=0.002), and COVID-related financial stress (β=0.088, interaction *p*=0.011), though not by pre-pandemic mood disorder (β=-0.048, interaction *p*=0.19). As shown in **Figure 2** and supported by stratified results (**Supplementary Table 3**), female participants and younger individuals had higher (two-to four-fold) predicted probabilities of depression than male participants and older individuals, respectively. In both cases, these effects appeared to attenuate at higher levels of social support. A similar pattern was observed for those reporting any pandemic-related financial stressors compared to those without any such stressors.

**Figure 2.**
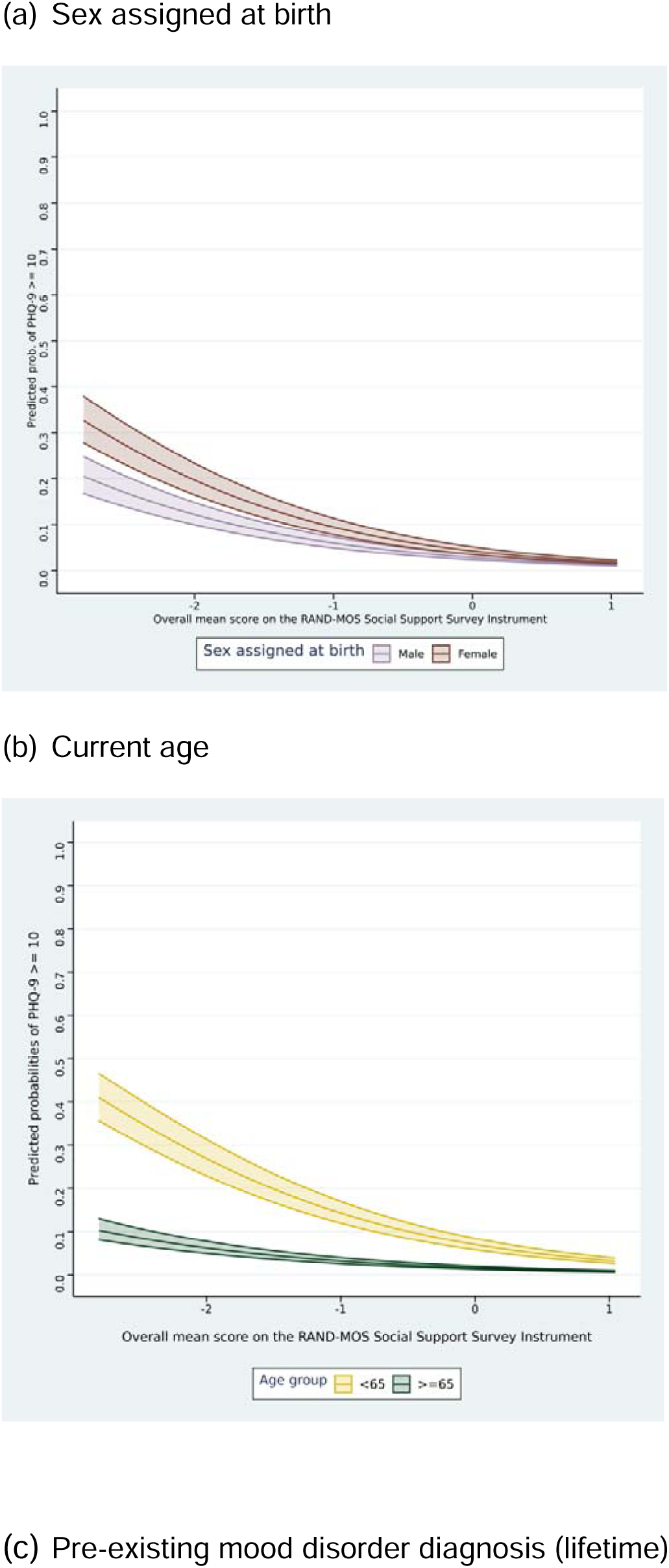
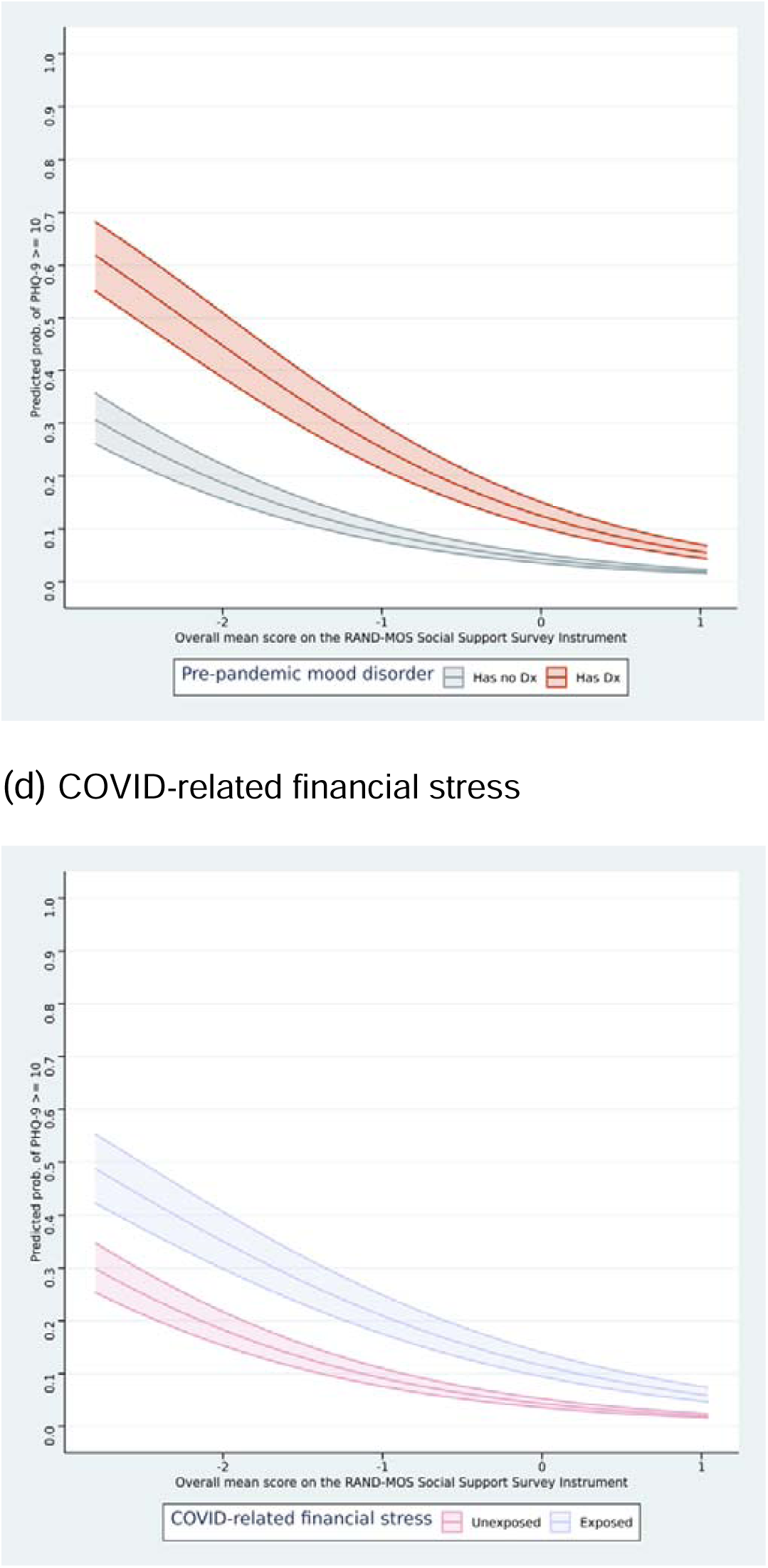
Variations in the predicted probabilities of depression at different levels of overall social support (standardized) by (a) sex assigned at birth, (b) current age, (c) pre-existing mood disorder diagnosis (lifetime), and (d) COVID-related financial stress.

## Discussion

In a large prospective cohort of 69,066 adults participating in the nationwide *All of Us* Research Program, higher levels of social support during the early months of the COVID-19 pandemic were associated with a 57% reduction in the odds of elevated depressive symptoms. Greater perceived support in multiple domains appeared most protective, with individuals reporting higher levels across tangible, emotional/informational, and positive social interaction supports showing a more than six-fold reduction in the odds of depression compared to those without. Moreover, female participants, younger individuals, and those reporting pandemic-related financial stressors showed generally increased odds of depression that were attenuated among those with higher levels of social support.

Among social support subtypes, examined both separately and in combination, emotional/informational support showed the largest protective association with depression, followed by positive social interactions, then tangible support. These findings are consistent with our prior analyses from the UK Biobank (16) showing that, among more than 100 potentially modifiable factors, confiding in others—which relates to the use and availability of emotional support—had the strongest protective association with incident depression in adults. This underscores the importance of trusted interpersonal outlets for mitigating depression risk, possibly via enhanced affect and cognitive regulation (17). Recent work in a longitudinal cohort found that perceived quality of one’s relationships was protectively associated with risk of psychiatric disorders during the pandemic (18), reinforcing the importance of social connection quality, not simply quantity. This is important to consider in the context of increased loneliness during the pandemic, which has also been linked to depression (19,20).

In addition to examining subtypes separately, we also probed the consequences of different subtype combinations of social support. In a dose-response fashion (**Figure 3**), compared to those reporting lower support on all three subtypes, individuals with higher support on all three subtypes showed the largest decrease in odds of depression, followed by those with higher support at least two subtypes, then those with higher support on one subtype alone. Most individuals endorsed support across all subtypes or none, though a sizeable subset of individuals (8.6%) endorsed higher tangible support alone. Tangible support was inversely associated with depression to a lesser degree compared to other subtypes. We hypothesized this association would have been stronger during the pandemic, given potential stressors of infection, illness, and quarantine that could incur practical challenges requiring assistance. Notably, we adjusted for demographic factors that could capture availability of tangible support in the home, including marital status, which may have attenuated observed effects.

**Figure 3.**
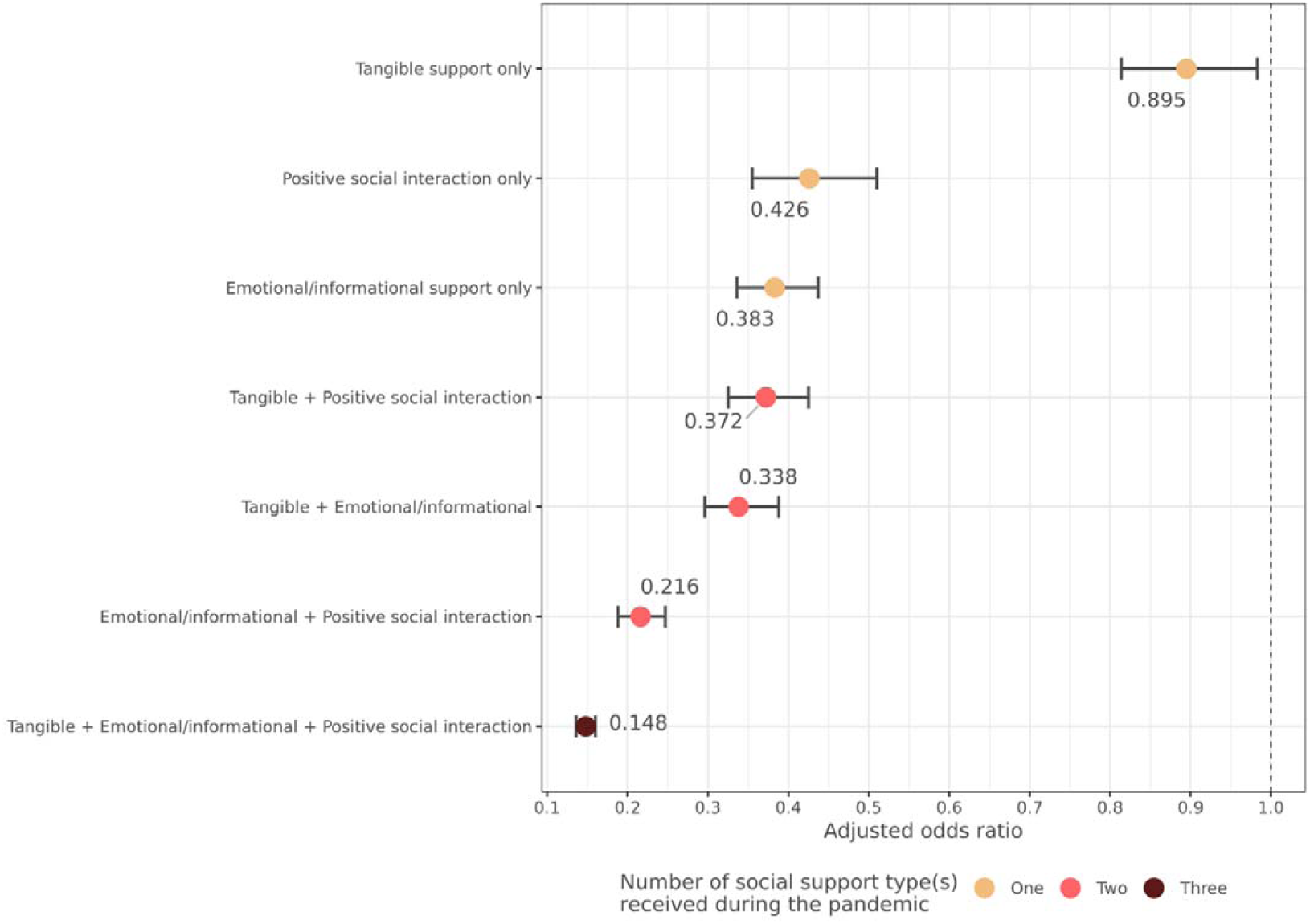
Odds ratio estimates from the mixed-effects logistic regression analysis of social support subtype combinations and depression risk.

When either higher emotional support or positive social interaction was added to tangible support, the odds of depression were further halved. Interestingly, higher emotional support and positive social interaction *together* seemed to show greater protective relevance for depression versus either subtype alone—dropping from a roughly 2.8-fold reduction to 4.6-fold reduction in depression odds—even in the absence of substantial tangible support. Endorsing higher positive social interaction alone was relatively rare (<2%), perhaps because time spent in positive experiences with others may naturally facilitate asking for tangible and/or emotional support; consistent with this, positive social interactions showed the highest correlations with both subtypes. Overall, results suggest that boosting social support on multiple fronts could produce the largest reductions in mental health risk. Social support-enhancing interventions often focus on the creation/use of support groups that provide a safe space for individuals to share concerns or interests (21,22) and may also include individual skill-building and network identification (23,24). Identifying which social support domains are perceived as high-quality versus lacking for a given individual may help in tailoring interventions.

Evidence of effect modification was observed for our studied intrinsic factors (i.e., age and sex) and during-pandemic factor (i.e., financial stressors) but not pre-pandemic factor (i.e., prior mood disorder diagnosis). Female participants generally had higher risk of elevated depressive symptoms compared to male participants, but showed larger reductions in depression odds in the presence of higher social support. This is consistent with a recent longitudinal study that also observed stronger protective associations between social participation/trust and any depressive symptoms during the pandemic among women versus men (25). Importantly, we expand prior work by identifying additional potential effect modifiers, including age and financial stressors related to the pandemic. Probing these results revealed that social support was linked to larger reductions in depression odds among participants at higher risk of elevated depressive symptoms during the pandemic (e.g., younger individuals, or those endorsing pandemic-related financial stressors). Thus, adults more vulnerable to depression—for intrinsic or environmental reasons—may benefit more from the protective effects of social support.

Strengths of this study included its longitudinal, multi-wave design in a large, diverse cohort with linked prior data and ongoing research participation, and nuanced operationalization of social support using an established survey instrument (7). While some studies may focus on the quantity of supportive individuals, studying perceived quality of support is an important feature of our research, as social support may be concentrated within a few individuals providing high-quality support. In addition, our study assesses the relationship of social support to clinically elevated symptoms of depression, which incur the greatest impairment and disability requiring prevention at the population level.

Several limitations should be noted. First, as with many survey instruments, our measure of social support captures self-reported perceptions of support, which may be influenced by concurrent depressed mood, thereby inflating the association. Notwithstanding, our primary analyses were consistent with lagged models in which baseline social support was associated with subsequent depression even after removing individuals with baseline elevated depression symptoms, suggesting the effect was not purely driven by contemporaneous mood states; however, further causal inference analyses are warranted (26). Second, we selected available variables from the COPE survey to examine their potential role as effect modifiers, but there may be other unmeasured intrinsic or pandemic-related factors (e.g., lockdown exposure) that may also influence the association between social support and depression. Our pre-pandemic mood disorder variable was also conservatively defined using linked EHR data which was available for most, but not all, participants. Third, given that most individuals were research volunteers with Internet access who were relatively well-educated and endorsed minimal financial stressors related to the pandemic, our sample may not generalize to more disadvantaged populations, though we used inverse-probability weighting as an attempt to match COPE survey participants more closely with the broader and more diverse *All of Us* Research Program study cohort.

## Conclusion

Social connection is increasingly recognized as a key public health priority (27). While the links between social support and depression are well established (2), a more nuanced understanding of this relationship—including which support subtypes are most relevant for depression risk, and who may benefit most, during a highly stressful global crisis—could inform targeted interventions to enhance resilience and reduce the population-level burden of depression. Individuals reporting higher levels of social support were at substantially reduced risk of elevated depressive symptoms during the COVID-19 pandemic. The perceived availability of emotional support and positive social interactions—and their combination—more so than tangible assistance, was key. Individuals more vulnerable to depression (e.g., women, younger individuals, and those experiencing financial stressors) may particularly benefit from enhanced social support during a major stressor, supporting a precision prevention approach.

## Data Availability

De-identified data are available on the Researcher Workbench of the *All of Us* Research Program located at https://workbench.researchallofus.org.

## Acknowledgements

The *All of Us* Research Program is supported by grants through the National Institutes of Health, Office of the Director: Regional Medical Centers: 1 OT2 OD026549; 1 OT2 OD026554; 1 OT2 OD026557; 1 OT2 OD026556; 1 OT2 OD026550; 1 OT2 OD026552; 1 OT2 OD026553; 1 OT2 OD026548; 1 OT2 OD026551; 1 OT2 OD026555; IAA#: AOD 16037; Federally Qualified Health Centers: HHSN 263201600085U; Data and Research Center: 5 U2C OD023196; Biobank: 1 U24 OD023121; The Participant Center: U24 OD023176; Participant Technology Systems Center: 1 U24 OD023163; Communications and Engagement: 3 OT2 OD023205; 3 OT2 OD023206; and Community Partners: 1 OT2 OD025277; 3 OT2 OD025315; 1 OT2 OD025337; 1 OT2 OD025276. In addition to the funded partners, the *All of Us Research Program* would not be possible without the contributions made by its participants.

All authors, except C.R.C., are supported by the International HundredK+ Cohorts Consortium (IHCC), a program of the Global Genomics Medicine Collaborative (GGMC) with support from the National Institute of Health, the Wellcome Trust, and the Chan-Zuckerberg Initiative. In addition, J.R.B. and S.B. are supported by: Dementias Platform UK (DPUK) funded by the Medical Research Council (MRC) MR/T0333771. J.W.S. is supported in part by a gift from the Demarest Lloyd, Jr. Foundation. K.W.C was supported in part by funding from the National Institute of Mental Health (K08MH127413) and a NARSAD Brain and Behavior Foundation Young Investigator Award.

