## Supplemental Table 1 for "Effects of social support on depression risk during the COVID-19 pandemic: What support types and for whom?"

### **Supplemental Table 1.** Definition of social support subtypes for the ten items from the RAND-MOS Social Support Survey Instrument.

| **Social support subtypes** | **Question** |
| --- | --- |
| Emotional/informational support | Someone to confide in or talk to about yourself or your problems. |
|  | Someone to turn to for suggestions about how to deal with a personal problem. |
|  | Someone who understands your problems. |
|  | Someone to love and make you feel wanted. |
| Positive social interaction | Someone to do things with to help you get your mind off things. |
|  | Someone to have a good time with. |
| Tangible support | Someone to take you to the doctor if you needed it. |
|  | Someone to prepare your meals if you were unable to do it yourself. |
|  | Someone to help with daily chores if you were sick. |
|  | Someone to help you if you were confined to bed. |
