## Supplemental Table 2 for "Effects of social support on depression risk during the COVID-19 pandemic: What support types and for whom?"

### **Supplemental Table 2**. Correlation matrix for the mean score of the three social support subtypes. Correlation coefficients were averaged across the three timings of survey administration.

| Pearson correlation (standard error) | Tangible | Emotional/informational | Positive social interaction |
| --- | --- | --- | --- |
| Tangible social support | - | - | - |
| Emotional/informational support | 0.657(0.002) | - | - |
| Positive social interaction | 0.702(0.002) | 0.828(0.002) | - |
