## Supplemental Table 3 for "Effects of social support on depression risk during the COVID-19 pandemic: What support types and for whom?"

### **Supplemental Table 3.** Stratified analysis examining the effects of overall social support on depression in each stratum of sex assigned at birth, pre-pandemic mood disorder diagnosis, and COVID-related financial stress.

| **Interaction term  being tested** | **Stratified results** | **Moderate to severe depression (PHQ-9 total score ≥ 10)** | | | |
| --- | --- | --- | --- | --- | --- |
|  |  | aOR | 95% CI | p-value | interaction p-value |
| Overall social support * sex assigned at birth | Male | 0.435 | (0.407 - 0.464) | <2.0E-16 | **1.7E-02** |
|  | Female | 0.433 | (0.416 - 0.449) | <2.0E-16 |  |
| Overall social support *  current age (2022) | Under 65 | 0.445 | (0.426 - 0.465) | <2.0E-16 | **2.2E-03** |
|  | 65 or older | 0.505 | (0.484 - 0.527) | <2.0E-16 |  |
| Overall social support *  pre-pandemic mood disorder dx | Without diagnosis | 0.441 | (0.425 - 0.457) | <2.0E-16 | 1.9E-01 |
|  | With diagnosis | 0.417 | (0.387 - 0.450) | <2.0E-16 |  |
| Overall social support *  COVID-related financial stress | Without COVID-related financial stress | 0.460 | (0.445 - 0.476) | <2.0E-16 | **1.1E-02** |
|  | With COVID-related financial stress | 0.386 | (0.340 - 0.439) | <2.0E-16 |  |

Abbreviations: aOR, adjusted odds ratio; CI, confidence interval; dx, diagnosis.

Note: All models were adjusted for sex assigned at birth, self-reported race, ethnicity, current age, marital/partnership status, homeownership, employment status, educational attainment, health insurance status, experience of COVID symptom(s), and diagnosis of mood disorder(s) within one year prior to the COVID-19 pandemic.
