## Supplemental Table 4 for "Effects of social support on depression risk during the COVID-19 pandemic: What support types and for whom?"

### **Supplemental Table 4.** Sex-stratified analysis examining the effects of overall and specific types of social support on depression.

| **Interaction term  being tested** | **Stratified results** | **Moderate to severe depression (PHQ-9 total score >= 10)** | | | |
| --- | --- | --- | --- | --- | --- |
|  |  | aOR | 95% CI | p-value | interaction p-value |
| Overall social support * sex assigned at birth | Male | 0.435 | (0.407 - 0.464) | <2.0E-16 | **1.7E-02** |
|  | Female | 0.433 | (0.416 - 0.449) | <2.0E-16 |  |
| Tangible social support * sex assigned at birth | Male | 0.646 | (0.607 - 0.687) | <2.0E-16 | **7.0E-02** |
|  | Female | 0.622 | (0.600 - 0.645) | <2.0E-16 |  |
| Emotional social support * sex assigned at birth | Male | 0.416 | (0.391 - 0.443) | <2.0E-16 | 1.2E-01 |
|  | Female | 0.422 | (0.407 - 0.437) | <2.0E-16 |  |
| Positive social interaction * sex assigned at birth | Male | 0.395 | (0.370 - 0.421) | <2.0E-16 | 7.2E-01 |
|  | Female | 0.433 | (0.418 - 0.449) | <2.0E-16 |  |

Abbreviations: aOR, adjusted odds ratio; CI, confidence interval; dx, diagnosis.

Note: All models were adjusted for sex assigned at birth, self-reported race, ethnicity, current age, marital/partnership status, homeownership, employment status, educational attainment, health insurance status, experience of COVID symptom(s), and diagnosis of mood disorder(s) within one year prior to the COVID-19 pandemic.
