## Supplemental Table 5 for "Effects of social support on depression risk during the COVID-19 pandemic: What support types and for whom?"

### **Supplemental Table 5.** Lagged analyses testing the association between baseline social support and depression risk assessed in either (a) the second wave only (1-month window) or (b) the second or third wave (2-month window), excluding individuals with elevated depressive symptoms at baseline

| **Outcome** | **N_respondent_** | **aOR** | **95% CI** | **p-value** |
| --- | --- | --- | --- | --- |
| Depression risk in June (1-month window) | 18,396 | 0.665 | [0.625, 0.709] | <2.0E-16 |
| Depression risk in June or July (2-month window) | 24,333 | 0.683 | [0.651, 0.716] | <2.0E-16 |

Note: All models were adjusted for sex assigned at birth, self-reported race, ethnicity, current age, marital/partnership status, homeownership, employment status, educational attainment, health insurance status, experience of COVID symptom(s) (measured in May of 2020), and diagnosis of mood disorder(s) within one year prior to the COVID-19 pandemic.
