## Supplemental Table 6 for "Effects of social support on depression risk during the COVID-19 pandemic: What support types and for whom?"

#### **Supplemental Table 6.** Descriptive statistics of the social support and depressive symptom scales.

(a) Distribution of mean item rating on the RAND-MOS social support scale by survey timing.

| **Survey timing** | **Mean** | **Median** | **SD** |
| --- | --- | --- | --- |
| May 2020 | 3.920 | 3.920 | 1.043 |
| June 2020 | 3.918 | 3.918 | 1.046 |
| July 2020 | 3.909 | 3.909 | 1.037 |

(b) Distribution of total sum score on the PHQ-9 scale by survey timing.

| **Survey timing** | **Mean** | **Median** | **SD** |
| --- | --- | --- | --- |
| May 2020 | 6.024 | 6.024 | 5.038 |
| June 2020 | 5.799 | 5.799 | 4.963 |
| July 2020 | 5.968 | 5.968 | 5.068 |
